# Structural Entropy of Daily Number of COVID-19 Related Fatalities

**DOI:** 10.1101/2020.10.19.20215673

**Authors:** Eren Unlu

## Abstract

A recently proposed temporal correlation-based-network framework applied on financial markets called Structural Entropy has prompted us to utilize it as a means of analysis for COVID-19 fatalities across countries. Our observation on the resemblance of volatility of fluctuations of daily novel coronavirus related number of deaths to the daily stock exchange returns suggests the applicability of this approach.

## I. Introduction

In December 2019, a novel coronavirus sourced atypical pneumonia cases are reported in Wuhan, China. Rapidly, it evolved in to an epidemic in its originating city [2]. Despite taken counter-measures, outbreak has gradually spread out globally, where World Health Organisation (WHO) declared it as a pandemic in March 11, 2020 and called for augmented enforcing policies to all governments [3]. Countries have responded to outbreak with varying degrees of containment and other preventive actions; also with varying latency [4] [5] [6] [7]. The pandemic is ongoing by the date September 20, 2020 when this paper is written, with worldwide 30 millions confirmed cases and one million deaths.

Authors in [1] have introduced a new correlation-based-network framework, where they refer as *Structured Entropy*. With their definition, this new framework is a revised interpretation of an earlier correlation-network method in the literature, *Structured Diversity*. Correlation-based-networks fall into a relatively well studied field of complex system analysis [8]. The algorithms of this category have been extensively investigated for a long time to model complex systems in diverse settings such as stock market dynamics, social networks, clustering human cerebral regions etc. [9] [10] [11] [1]. We can see that most of the publications focus on the dynamics of financial data, where nodes are represented as assets such as a stock price of a company or a certain currency rate. The central idea is to connect the similar nodes within certain time periods which show high correlation [10]. [1] underlines the fact that these methods lack continuous observation, which is a vital aspect in the analysis of financial market.

Therefore, they overcome this issue with their newly provided Structured Entropy, where correlations between assets (nodes) are measured continuously in a pre-defined long sliding window. Rather than using correlation matrices per window, the authors prefer to use *community based networking*, where the nodes are assigned to a community (cluster) in a one-to-one and binary fashion by thresholding correlation values. After constructing communities for each time window, a diversity measure per window is calculated, where [1] calls as *Structural Entropy*, which itself is inspired by the concept of *Shannon index* [12]. This scalar measure takes the number of communities and their size; where it offers a more fine grained representation of the actual dynamics according to authors’ claim.

It is a repeatedly proven hypothesis under numerous different market conditions that short term fluctuations of asset values such as stock prices or currency rates can be modeled well with a Gaussian distribution [11]. This highly volatile nature makes short term algorithmic trading one of the most challenging tasks for machine learning. [1] successfully explain the overall volatility and heterogeneity of the stock market continuously with this technique, while temporally clustering the correlated companies.

We have observed that daily number of COVID-19 fatalities in a country follow a very similar highly volatile pattern, increasing-decreasing arbitrarily, just as daily stock market returns. This observation has prompted us to apply the Structured Entropy method to explain the volatility and heterogeneity of the daily number of deaths among countries since the beginning of the pandemic.

## II. Structured Entropy

Structured Entropy, a single scalar value on a specific time step defines the actual heterogeneity of the global system. It is calculated on each time step independently on two stages. First, the nodes are assigned to communities, such that each node can only be a member of a single community. Isolated nodes constitute their very own single member clusters. For this purpose, [1] slides a pre-defined length window on each time step over the values of each node, and calculates the overall cross-correlation matrix of the nodes. In other words, the cross-correlation matrix at a step is calculated based on the *τ* previous values of that step, *τ* being the window length (Eq. 1) [1].

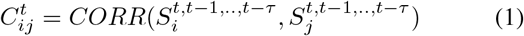

Based on this correlation matrix, a binary adjency matrix is constructed which defines the affiliation of each member to a community. Researchers use a simple thresholding mechanism, where if the correlation of two nodes are higher than a threshold, they are assigned to the same cluster. However, authors carefully highlight the possibility of utilization of Random Matrix Theory based methods for more sophisticated clustering. For the sake of the integrity, we also follow the same approach in this work. However, we have preferred to determine the optimal threshold for correlation matrix based clustering by searching for the value which yields the largest standard deviation of temporally changing structured entropy, as basically variance is correlated to the overall information encapsulated in a system.

After each node is assigned to a cluster, the instantaneous Structural Entropy, the measure of heterogeneity is calculated based on the number of communities and community sizes as follows :

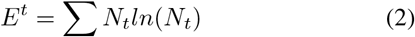

where *N*_*t*_ is the number of communities at the time.

## III. COVID-19 Related Daily Number of Fatalities

As mentioned previously, we have made the observation that the daily number of COVID-19 related deaths show similar high volatility patterns just like the well known log returns of daily stock prices. We have chosen 26 countries where total number of confirmed COVID-19 related deaths has exceeded 5,000. The dataset comprises the daily fatalities in each of these 26 countries since March 15, 2020; when relatively we start to observe to have statistically enough data. Fig. 1 illustrates the overall correlation matrix and Gaussian like distribution (with considerable heavy-tail effect) of daily percentage of change of log-fatalities of 26 countries since March 15, 2020. And Fig. 2 includes two time series graphs for average and standard deviation of daily sliding window (20 days) percentage of change of log-fatalities percentage of change of 26 countries since March 15, 2020. The red line indicates the overall of all 26 countries.

**Fig. 1.**
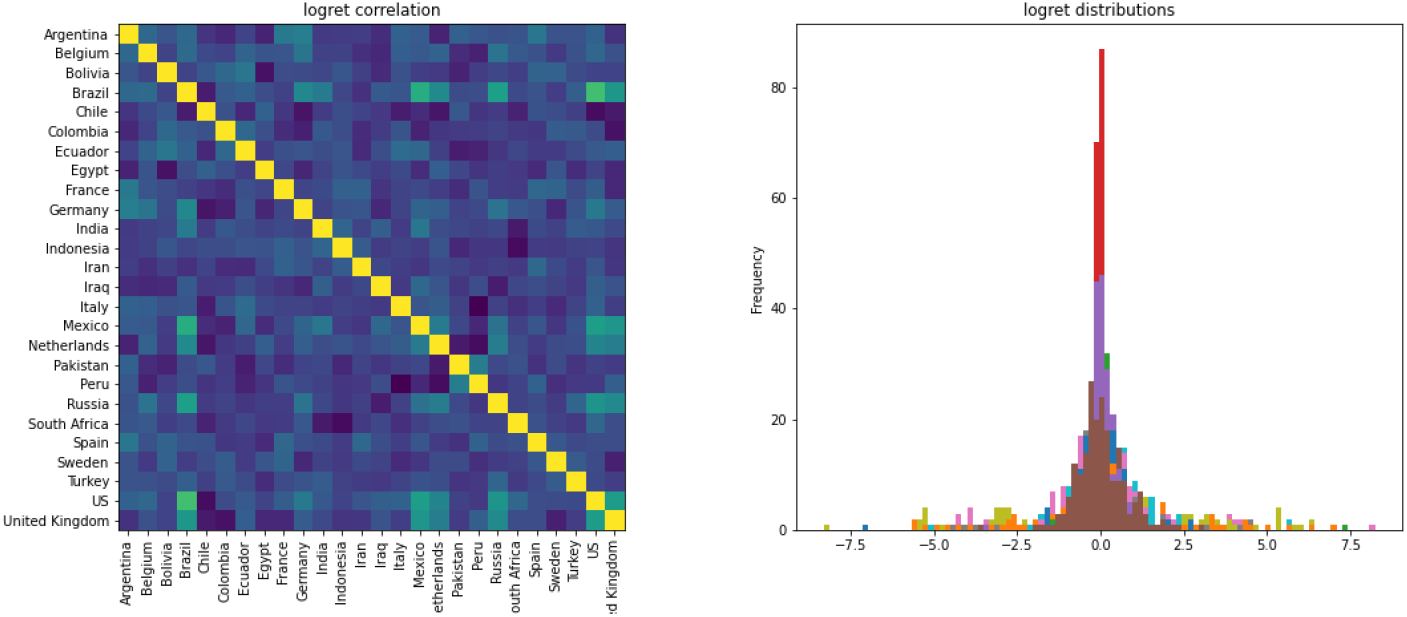
Overall correlation matrix and Gaussian like distribution (with considerable heavy-tail effect) of daily percentage of change of log-fatalities of 26 countries since March 15, 2020.

**Fig. 2.**
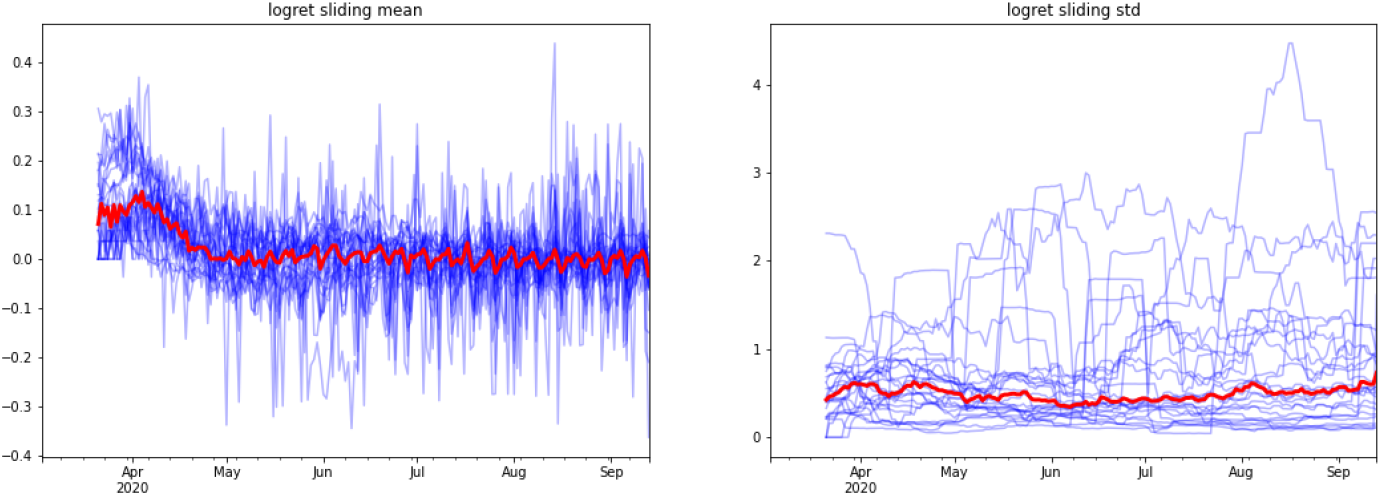
Average and standard deviation of daily sliding window (20 days) log-fatalities percentage of change of 26 countries since March 15, 2020. The red line indicates the overall of all 26 countries.

The choice for the window is a matter of important discussion in these types of settings. One shall also approach the optimal window selection issue considering the attributes and dynamics of the system. Taking into account the observed COVID-19 epidemiological statistics such as average number of days it takes to reach since the first contact with the virus to the infectiousness, hospitalisation, fatality states etc. [13] [14], we have determined the window length as 20 days.

As mentioned previously, we have determined the most informative correlation matrix threshold by a brute force search for maximizing the variance of temporally changing structured entropy of the overall system, where we base our claim on the rationale that the retained information is correlated to the variance.

Fig. 3 illustrates the evolution of instantaneous structured entropy value since March 15, 2020 for different correlation matrix threshold values. We have plotted the standard deviation of overall structured entropy with varying community affiliation threshold on correlation matrices in Fig. 4. 0.475 gives the largest standard deviation, suggesting this value shall preserve the most of the information, thus we have used this threshold for our work.

**Fig. 3.**
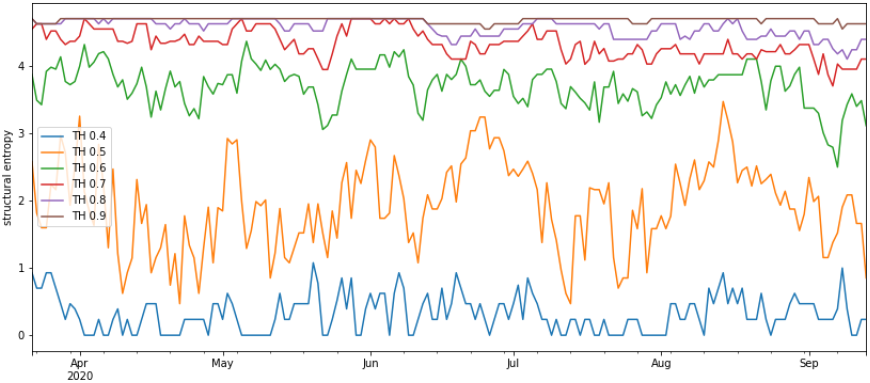
Structured entropy for different community affiliation thresholds on correlation matrices.

**Fig. 4.**
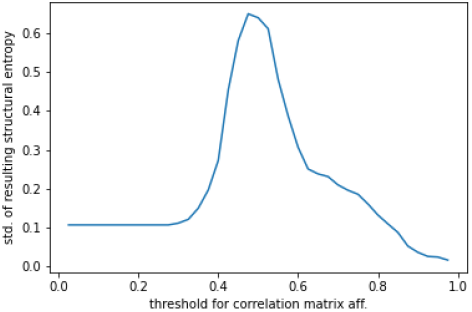
The standard deviation of overall structured entropy with varying community affiliation threshold on correlation matrices. 0.475 gives the largest standard deviation, suggesting this value shall preserve the most of the information, thus we have used this threshold for our work.

**Fig. 5.**
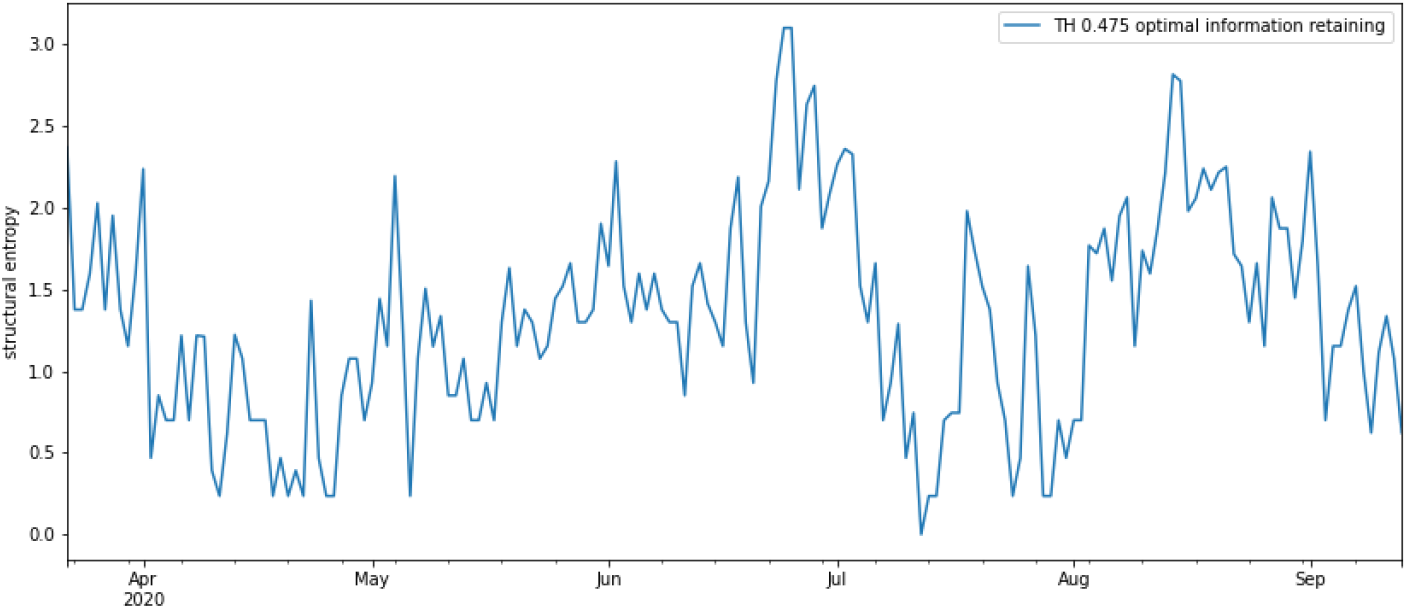
Structured entropy of daily log percentage of change of fatalities of 26 countries since March 15, 2020 (window of 20 days) with community affiliation threshold of 0.475.

Next, we show the evolution of daily structured entropy value with the threshold of 0.475 since March 15, 2020 in Fig. 6. As it can be seen there is a sudden peak and a following drastic fall between June 15, 2020 and first week of the july. This suggests that around this time countries started to show very diverse patterns in number of daily fatalities, and starting from july the overall scheme started to converge to a similar process. This shall stem from the outlying few countries who shows more unpredictable patterns. However, as it can be seen from Fig. 6 provides us a compact and informative graph to evaluate the behaviour of major countries.

**Fig. 6.**
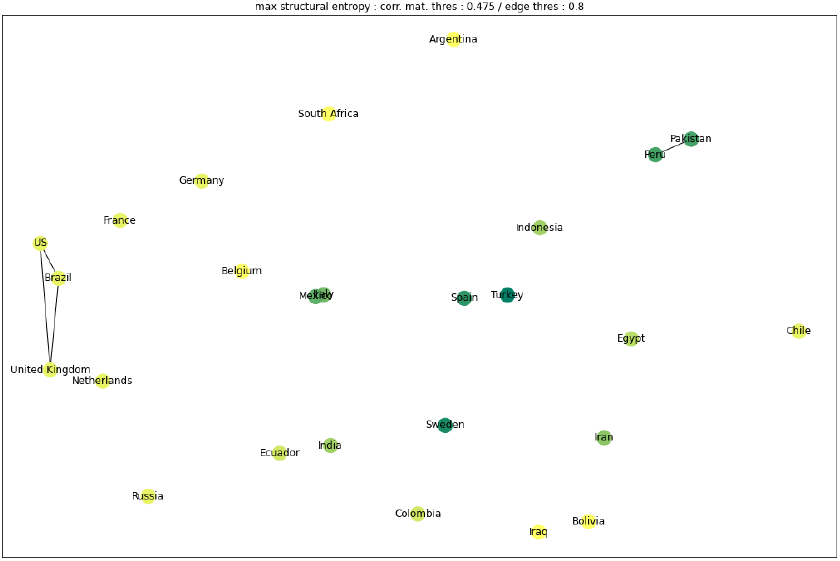
The clustering of countries on the day with the maximum entropy with a relatively large edge strength threshold of 0.8. We see that almost all countries show *sui generis* process for daily number of patters in last 20 days, except for 2 formed communities. Peru and Pakistan show an instantaneous resemblance. Interestingly, United States, Brazil and United Kingdom form a cluster, which are countries sparked discussions due to their controversial counter-epidemic measures.

**Fig. 7.**
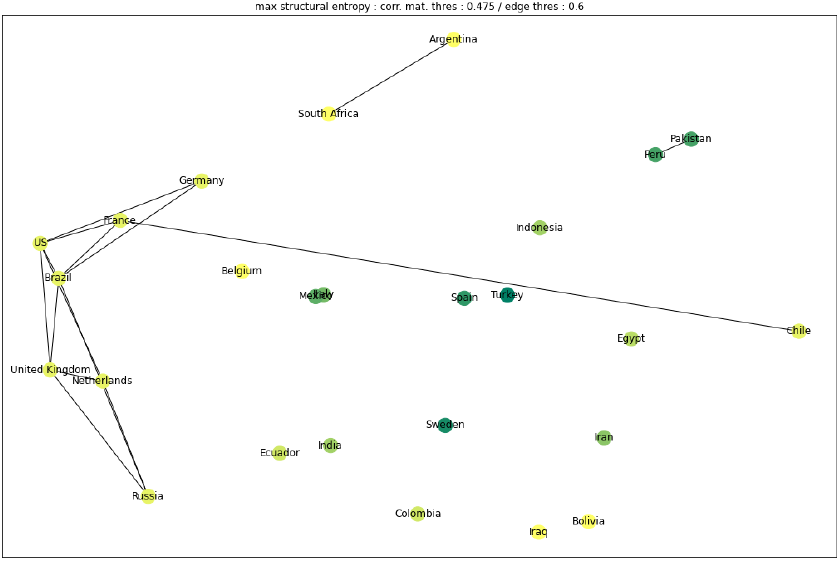
The clustering of countries on the day with the maximum entropy with a relatively large edge strength threshold of 0.6. As we decrease the threshold of edge formation, we see that Russia, Netherlands, Germany and Chile joins the community of United States, Brazil and United Kingdom. Note that maximum entropy is observed on June 24, 2020.

**Fig. 8.**
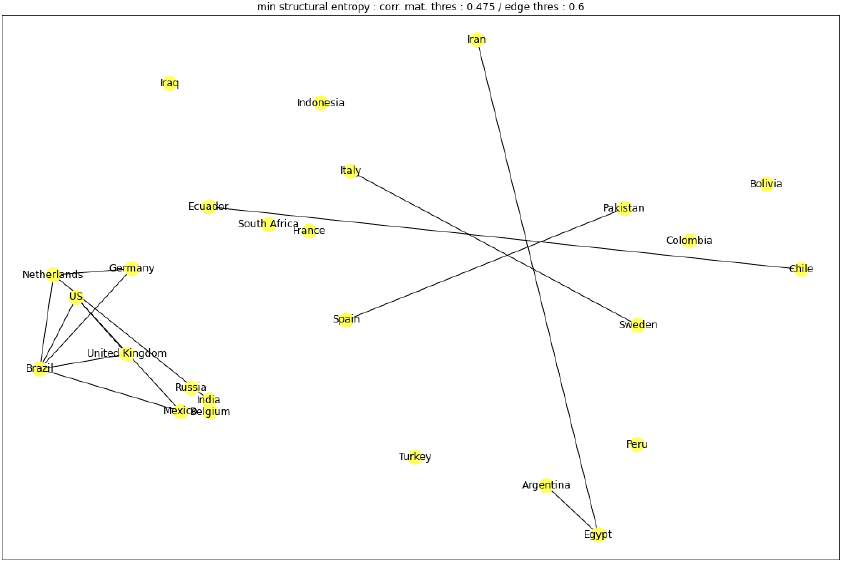
The clustering of countries on the day with the minimum entropy with edge strength threshold of 0.6. We observe a large cluster formed by mostly European countries. Note that minimum entropy is observed on July 8, 2020.

Next, we investigate the clustering of countries for different times with different structured entropy values. For instance, we evaluate the instantaneous clustering on maximum, minimum and overall entropy for different edging (node connection strength in a graph). As [1], for this purpose we first calculate the 2 component Principal Component Analysis (PCA) values of the correlation matrices to reduce the setting to a two dimensional representation before constructing graphs.

## IV. Conclusion

In this paper, we have applied the recent noteworthy framework called Structured Entropy which has been used for analyzing stock markets to the daily number of COVID-19 related deaths in various countries. The inspiration stems from our observation on the highly volatile fluctuations of daily number of fatalities in countries. We have demonstrated that structured entropy concept, and possibly other types of correlation-based networking algorithms can aid significantly policy makers and analysts in an epidemiological context.

## Data Availability

Data are public coronavirus statistics

## Acknowledgements

We thank Assaf Almog and Erez Shumeli for providing their source codes online of their seminal paper [1], which is the central idea applied in this work.

## Notes

### Competing Interest Statement

The authors have declared no competing interest.

### Funding Statement

No funding

